# Integrating Preconception Risk Assessment and Counseling with Primary Health Care: A Feasibility Assessment

**DOI:** 10.1101/2024.04.17.24305399

**Authors:** Anne L. Dunlop, Susana Alfonso, Nora Hansen, Dionne Williams, Ariela L. Marshall, Victoria M. Anderson

## Abstract

**Background:** Professional association recommendations call for integrating preconception health promotion with primary care, yet there are scarce tools and implementation research to support practices in doing so.

**Purpose:** To evaluate the feasibility of integrating a preconception health digital risk assessment and virtual coaching into women’s primary care encounters.

**Methods:** In the Emory Family Medicine Clinic (Atlanta, Georgia), female patients 21-40 years of age with a well-woman or chronic condition encounter scheduled between 9/1/2022 and 5/1/2023 were invited to participate. Consenting patients were provided the *Frame Your Future* weblink to complete the digital risk assessment followed by virtual counseling, and their family physicians were provided with a pdf summary to discuss during the primary care encounter. Demographic and clinical information was collected via medical record abstraction and patient and physician experiences via survey.

**Results:** Of 46 enrolled patients, 44 (96%) made a Frame account, 38 (86%) completed the risk assessment, 34 (89%) completed virtual coaching, and 24 (71%) had a physician discuss their preconception health assessment during the primary care encounter. Nearly 80% of patients reported an increase in confidence in discussing fertility with their physician, and 50% reported they would not otherwise have brought up fertility and preconception health during the encounter. Both patients and physicians were satisfied with the process and viewed it as helping motivate positive changes in patient health and health behaviors.

**Conclusion:** The completion of preconception digital risk assessment and virtual counseling facilitates discussion of preconception health during primary care encounters and shows promise for improving women’s health.

## Introduction

The Centers for Disease Control and Prevention (CDC) and the American College of Obstetrics and Gynecology (ACOG) recommend preconception health promotion for all persons of reproductive capability and age.^1,2^ In 2006, a Select Panel recommended the provision of preconception health promotion (risk assessment, health education and risk-specific counseling) *as part of primary care* for all individuals capable of becoming pregnant with the goal of improving health and reproductive outcomes (to include having desired, well-timed pregnancies that end in healthy maternal and infant outcomes).^3^ Although patients express interest in preconception counseling, less than 14% of US ambulatory visits include any.^4–7^ Competing demands for health care provider attention along with lack of knowledge and consensus on the screening and counseling approach, and failure to recognize all individuals for whom services are indicated (*e.g.,* those who are unmarried, in same-sex relationships, gender diverse) are barriers to the broad provision of preconception health promotion.^8–11^ Those who receive care in community primary care clinics, rather than in dedicated women’s health settings, are particularly affected by these barriers,^12^ emphasizing the importance of identifying strategies to systematically implement such services in clinics that serve low-access populations.^13,14^ Despite these recommendations, calls to action, and evidence based guidelines, there is little health services research to guide implementation.^15,16^

To fill this gap, Frame in partnership with clinicians with expertise in reproductive health created a preconception health risk assessment and counseling package that includes a standardized web-based screening questionnaire (with embedded skip logic and conditional branching) followed by a virtual coaching session. Since 2020, individuals have been able to use the Frame website (www.frameyourfuture.com) to create an account, complete the digital risk assessment, and access health education and coaching without a physician. Frame has been evaluated for client acceptability as both a direct-to-consumer assessment tool and as a tool integrated with health care provider assessments in fertility, gynecology, and women’s health clinics.

### Purpose

To evaluate the feasibility and acceptability of integrating Frame’s preconception health digital risk assessment and coaching package into primary health care encounters at a family medicine clinic.

## Methods

### Design Overview

Eligible participants were identified through chart review of patients scheduled with participating family physicians between 9/1/2022 and 5/1/2023. Consenting patients were asked to complete the Frame digital risk assessment and a virtual health coaching session prior to their primary care encounter, and their physicians were provided a pdf summary of identified preconception and fertility risks and notes from the virtual coaching session to facilitate provision of preconception health promotion during the primary care encounter. Patient participants were asked to complete surveys immediately and one month after the virtual coaching and immediately following the primary care encounter. Participating physicians were also asked to complete a survey immediately following the primary care encounter. Medical record abstraction was used to collect participant demographic information and clinical characteristics. This study was reviewed and approved by the Institutional Review Board of Emory University; written informed consent was obtained from research participants.

### Setting

This study was conducted in Emory Family Medicine Clinic in Dunwoody, Georgia, a residency training site that serves patients of diverse ages, educational levels, genders and insurance status.

### Participants

Eligibility for patient participation included: (1) Assigned female at birth (regardless of gender identity); (2) Between 21-40 years of age; (3) English-speaking (as Frame is available in only English); (4) Scheduled for an annual wellness and/or a follow-up for a chronic health condition with physicians who agreed to take part in the study (two faculty, five family medicine residents); (5) Not pregnant, diagnosed with infertility, or have undergone sterilization at the time of enrollment; (6) Capacity to consent.

### Procedures

After review of scheduled patients, those who appeared eligible were contacted by a research team member via phone to explain the study, complete eligibility screening and informed consent.Those who consented were e-mailed a link to the Frame web portal, which they could use to create a profile, complete the digital risk assessment, and authorize their physician to access their report. Participants were then able to schedule a virtual coaching session with a Frame health coach after which the physician’s team was provided with a link to a Physician Dashboard with a pdf summary; clinical staff also downloaded this pdf, provided a hard copy to the physician during the scheduled encounter, used the “sticky note” function in the electronic health record to indicate that the summary was available for the encounter.

### Data Collection

#### Patients

Prior to the digital assessment, participants completed a survey adapted from the 5-item Perceived Efficacy of Patient-Physician Interaction (PEPPI) to measure baseline confidence in talking with their provider about fertility and understanding factors that impact fertility. Following coaching, participants were e-mailed links to repeat the PEPPI (to assess for change) and to complete a 2-item survey to assess satisfaction with the coaching. During the month after coaching, participants were offered asynchronous coaching support via e-mail and text and at the end of the month, satisfaction with coaching and health status was assessed via repeat survey. After the primary care encounter, participants were e-mailed a 7-item survey to assess understanding of next steps, impressions of whether the digital assessment and coaching faciliated conversations with the provider, intended and undertaken health and health care behaviors, and needed information and supports. Patients were provided two $25 electronic gift cards for completion of the one-month post-coaching and the post-encounter surveys. The research team completed medical record abstraction to gather sociodemographic and clinical data.

#### Physicians

Following the primary care encounter, participating physicians were sent a link to an 8-item survey to capture whether the Frame assessment was discussed and, if so, perceptions regarding whether its discussion facilitated aspects of the encounter or changes in patient care.

### Data Analysis

Using collected data, we assessed: (1) Rates of participant completion of the digital risk assessment and having a review of the assessment during the primary care encounter, and whether there were differences by sociodemographic (age group: 21-29, 30-40; insurance status: government, private; education level: high school or less, some college or more) or clinical characteristics (presence of chronic health conditions) using Student’s t-test or Wilcoxon test (for continuous measures) and Chi-square test or Fisher’s exact test (for categorical measures), as appropriate for the data. (2) Change in confidence and health literacy via tabulating and comparing responses to confidence and literacy questions prior to and immediately following coaching. (3) Patient satisfaction with and perceived utility of Frame by tabulating the patient post-coaching and post-primary care encounter questionnaires. (4) Primary health care provider satisfaction with and perceived utility of Frame was by tabulating physician responses to questionnaires completed after each encounter with an enrolled participant. (5) The needed inputs for implementation of Frame into primary health care by collecting coordinator and health care provider feedback regarding perceived facilitators and barriers.

## Results

During the recruitment period of 9/1/2022 through 5/1/2023, 214 (85%) of 251 eligible patients were contacted by the research team; the remainder could not be reached. Of these, 89 patients (42%) expressed interest in the study and were e-mailed the consent form. Of those expressing interest, 46 (52%) consented and were enrolled. Of the 46 enrollees, 44 (96%) made a Frame account, 38 (86%) completed the digital risk assessment and 34 (89%) completed the virtual health coaching. Of those completing coaching, 24 (71%) had their assessment discussed during a primary care encounter (**Figure 1**). Among the 10 who completed coaching but did not have it discussed: 4 had rescheduled their appointment such that it occurred prior to their coaching session, 5 canceled their appointment, and 1 missed their appointment. Among the 34 participants who completed coaching, 31 (91%) engaged with their health coach via text or e-mail at least once during the month following.

**Figure 1.**
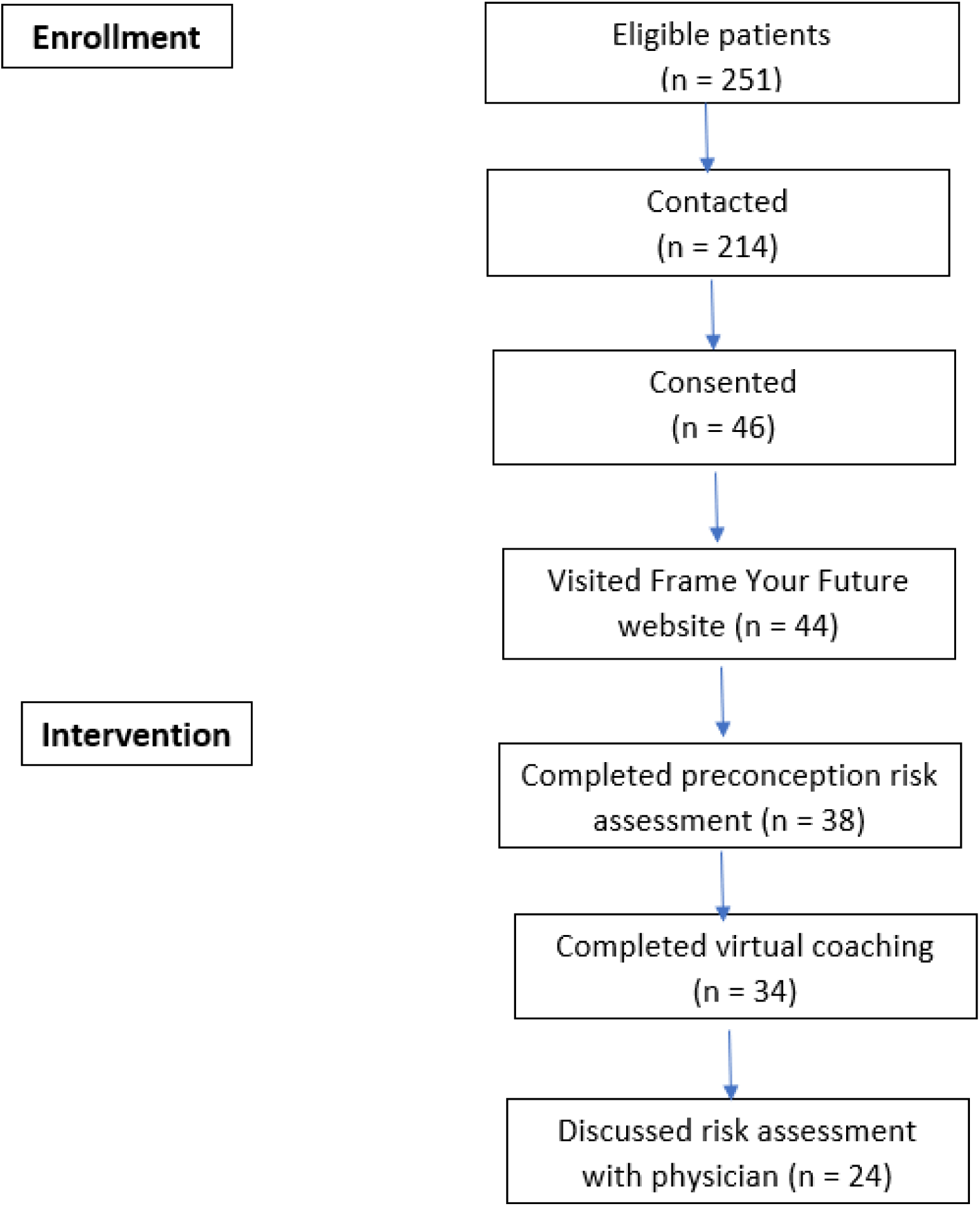
Participant recruitment flowchart.

Sociodemographic and clinical characteristics of the 46 consenting participants (overall and according to whether they completed coaching and discussion of the assessment with their physician) are given in **Table 1**. Approximately two-thirds were between 30 and 40 years of age, three-quarters had some college or higher education, half were non-Hispanic Black, one quarter had government insurance, two-thirds had no prior birth, 80% had a diagnosis of one or more chronic medical conditions and almost half used a prescription medication regularly. There were significant differences in the percentage who completed the virtual coaching session by patient race-ethnicity (52% for non-Hispanic Black patients, 86% for Hispanic patients, 92% for white patients) and parity (84% and 50% for those without and with a prior birth, respectively). There were also significant differences in the percentage who had their assessment discussed with their physician by insurance type (33% and 62% for those with public and private insurance, respectively) and parity (63% and 29% for those without and with a prior birth, respectively).

**Table 1.** Sociodemographic and Clinical Characteristics of Participants Overall and by Completion of Frame Virtual Coaching and Discussion of Assessment with Physician.

| Characteristic | All<br>Participants<br>N = 46 | Completed Virtual Coaching |  |  | Discussed Assessment with Physician |  |  |
| --- | --- | --- | --- | --- | --- | --- | --- |
|  |  | Yes<br>N = 34 | No<br>N = 12 | p-value | Yes<br>N = 24 | No<br>N = 22 | p-value |
| Age in years |  |  |  |  |  |  |  |
| Mean age +/- sd | 31.5 ± 5.9 | 31.4 ± 6.0 | 31.7 ± 5.8 | 0.911 | 32.1 ± 5.9 | 30.9 ± 6.0 | 0.489 |
| Age group |  |  |  |  |  |  |  |
| 21-29 y | 17 (37%) | 14 (82%) | 3 (18%) | 0.489 | 12 (71%) | 5 (29%) | 0.053 |
| 30-40 y | 29 (63%) | 20 (69%) | 9 (31%) |  | 12 (41%) | 17 (59%) |  |
| Education level |  |  |  |  |  |  |  |
| High school or less | 11 (24%) | 7 (64%) | 4 (36%) | 0.425 | 3 (27%) | 8 (73%) | 0.055 |
| Some college or more | 33 (72%) | 26 (79%) | 7 (21%) |  | 20 (61%) | 13 (39%) |  |
| Missing | 2 (4%) |  |  |  |  |  |  |
| Racial/ethnic group |  |  |  |  |  |  |  |
| Non-Hispanic Black | 21 (46%) | 11 (52%) | 10 (48%) | 0.027* | 10 (48%) | 11 (52%) | 0.668 |
| Non-Hispanic White | 13 (28%) | 12 (92%) | 1 (8%) |  | 7 (54%) | 6 (46%) |  |
| Non-Hispanic Other | 5 (11%) | 5 (100%) | 0 (0%) |  | 4 (80%) | 1 (20%) |  |
| Hispanic, Any race | 7 (15%) | 6 (86%) | 1 (14%) |  | 3 (42%) | 4 (58%) |  |
| Insurance type |  |  |  |  |  |  |  |
| Public | 12 (27%) | 6 (50%) | 5 (50%) | 0.052 | 3 (33%) | 9 (66%) | 0.044* |
| Private | 34 (73%) | 28 (82%) | 6 (18%) |  | 21 (62%) | 13 (38%) |  |
| Prior births |  |  |  |  |  |  |  |
| None | 32 (70%) | 27 (84%) | 5 (16%) | 0.027* | 20 (63%) | 12 (17%) | 0.034* |
| One or more | 14 (30%) | 7 (50%) | 7 (50%) |  | 4 (29%) | 10 (71%) |  |
| Chronic conditions |  |  |  |  |  |  |  |
| Mean number ± sd | 2.0 ± 1.5 | 2.2 ± 1.5 | 1.3 ± 1.3 | 0.069 | 2.3 ± 1.6 | 1.6 ± 1.3 | 0.093 |
| Any chronic condition |  |  |  |  |  |  |  |
| Yes | 37 (80%) | 29 (78%) | 8 (12%) | 0.211 | 21 (57%) | 16 (43%) | 0.276 |
| No | 9 (20%) | 5 (56%) | 4 (44%) |  | 3 (33%) | 6 (66%) |  |
| Prescription medications |  |  |  |  |  |  |  |
| Yes | 22 (48%) | 17 (77%) | 5 (23%) | 0.619 | 12 (55%) | 10 (45%) | 0.758 |
| No | 24 (52%) | 17 (71%) | 7 (29%) |  | 12 (50%) | 12 (50%) |  |

Of the 34 participating patients who completed virtual coaching, 29 completed both a baseline and post-coaching confidence and literacy survey; of these, 79% showed an increase in confidence in talking about fertility with a health care provider and 66% showed an increase in confidence around understanding factors that impact their fertility (**Table 2**).

**Table 2.** Participant Confidence and Literacy around Discussing Fertility Health – From Baseline to Immediately Post-Coaching (29 respondents)

|  |  |
| --- | --- |
| <u>Question:</u> *How confident are you in talking with your provider about your fertility on a scale of 1-10 with 1 being not at all confident and 10 being very confident?* |  |
| Baseline score = 6.4 | Post-coaching score = 9.2 |
| 23 (79%) with pre-post increase in confidence score<br>43% average increase in confidence score |  |
| <u>Question:</u> *How confident are you in understanding factors that impact your fertility health on a scale of 1-10 with 1 being not at all confident and 10 being very confident?* |  |
| Baseline score = 5.9 | Post-coaching score = 8.4 |
| 19 (66%) with pre-post increase in confidence score<br>43% average increase in confidence score |  |

Fourteen participants completed the immediate post-coaching survey; of these, 13 reporting being ‘Very Satisfied’ and 1 reporting being ‘Satisfied’. **Table 3** shows narrative responses of patients’ perceptions of gaining knowledge from their coach, connecting with their coach on a personal level, and being linked to resources or motivated into action by their coach. Of the 31 participants who completed the one-month post-coaching survey, 22 (71%) reported that their health had improved (2 significantly, 5 moderately, 15 slightly).

**Table 3.** Patient Narrative Responses Post-Coaching, Organized by Theme.

|  |
| --- |
| **THEME: Gain in Knowledge** |
| My coach was very knowledgeable. |
| My coach gave me good advice. |
| She offered real time, attainable information and suggestions. |
| I was able to learn many new things and have my questions answered. |
| During the coaching session, a lot of factors were explained and my questions answered. The process was thoroughly explained and I understand the following next steps. |
| I felt incredibly informed. |
| **THEME: Connected on Personal Level** |
| My coach and seemed to understand some of the concerns I had. |
| My coach is the best! She was excellent in creating a warm, informative, non-judgmental environment for me to share my reproductive goals. We also connected on a cultural level, which I appreciated and valued. |
| She is very attentive, helpful, and willing to do things in an alternate format (text and phone instead of e-mail). |
| I think my coach was a great coach with a lovely attitude and very personable. |
| I felt heard and respected. |
| **THEME: Linked with Resources or Motivated Action** |
| I am satisfied with the work my coach and I did together because she was very detailed and thought outside the box. What was especially nice was although we spoke about women's health, I was also able to talk about other health concerns of mine. My coach was even able to assist me in finding a therapist to help me work through some mental health struggles as well. |
| I felt empowered after our call and also called to make some steps toward making space in my life for expanding my family in the future if I choose to do so. |

Of the 24 participants who completed the post-primary care encounter survey, 50% indicated they would not have brought up their fertility during the physician encounter otherwise, whereas 3 (12%) were unsure and 9 (38%) indicated they would have brought up fertility issues anyhow. Nearly all (11 of 12) of those who would not have brought up fertility and all (3) who were ‘unsure’, had at least chronic medical conditions; this set of 15 patients received lifestyle counseling (6), new diagnoses (2), referrals for care (2), changes in medications (2) or contraceptives (2), discussed changes in timing of planned childbearing (4), and received vaccinations (2). Two patients with multiple chronic conditions asked about why fertility preservation and reproductive health maximization had not been addressed in previous primary care visits to the practice. A substantial percentage (20, 83%) reported they made specific changes as a result of the encounter (**Table 4**), with the most common changes being lifestyle related such as increasing physical activity (11), reducing alcohol (3), marijuana (2), and tobacco (1).

**Table 4.** Patient-reported Activities or Change Following Discussion of Frame with Physician.

| Activity or Change | Number (%) |
| --- | --- |
| Improved lifestyle | 13 (54%) |
| ▪ Increased physical activity | 11 |
| ▪ Improved diet | 10 |
| ▪ Plan to lose weight | 9 |
| ▪ Reduce alcohol | 3 |
| ▪ Reduce marijuana | 2 |
| ▪ Reduce tobacco | 1 |
| Began taking prenatal vitamin | 10 (42%) |
| Looked into benefits and coverage | 10 (42%) |
| Had conversation with partner | 8 (33%) |
| Scheduled additional physician visit | 8 (33%) |
| Adjusted medications | 7 (29%) |
| Adjusted birth control | 7 (29%) |
| Changed timing of planned childbearing | 6 (25%) |
| Began tracking ovulation | 5 (21%) |
| Pursued additional fertility testing or work-up | 4 (17%) |
| Discussed chronic condition management with provider | 3 (12%) |
| Began trying to conceive | 3 (12%) |
| Pursued egg preservation | 1 (4%) |

Physician survey responses are given in **Table 5**. Of the 23 surveys completed, none indicated disagreement with any of the questions about the utility of Frame. Overall, physician survey responses had a favorable impression, with the following strongly agreeing or agreeing that Frame: made the patient more engaged in care (20, 88%); helped the patient be healthier, better informed, and enhanced quality of the visit (19, 83%); made the visit more efficient (17, 80%); helped provide the best care (16, 70%); and improved clinical outcomes (11, 48%). All (100%) surveys indicated that a specific care activity occurred during the encounter as a result of discussion of the Frame assessment, including: discussion of lifestyle modifications (12, 52%), with topics of counseling including obesity and overweight (5 encounters), excessive alcohol use (2 encounters), marijuana use (3 encounters), and tobacco use (2 encounters). Other specific care activities included: ordering laboratory studies or evaluations to address symptoms or concerns for 6 encounters (26%), including hormone or endocrine testing for irregular cycles, following up on past diagnoses of iron-deficiency anemia and pre-diabetes; discussing the timing of childbearing for 5 encounters (22%); diagnosing a new condition or changing medications (to a medication safer in pregnancy among those planning to have a child in the near future) which was documented for 4 encounters (17%); changing contraceptive method to be more compatible with their reproductive desires, referring the patient to another type of provider (such as an obstetrician-gynecologist, fertility specialist, geneticist, or social worker), as well as providing indicated vaccines, each of which was documented for 3 encounters (13%).

**Table 5.** Physician Responses About Extent of Agreement with Statements About Frame.

| Statement | Strongly Agree | Agree | Neither Agree/Disagree | Disagree | Strongly Disagree |
| --- | --- | --- | --- | --- | --- |
| Helped me provide the best care possible | 7 (30%) | 9 (40%) | 7 (30%) | 0 | 0 |
| Made the visit more efficient | 4 (23%) | 13 (57%) | 6 (26%) | 0 | 0 |
| Made my patient more engaged in care | 9 (39%) | 11 (49%) | 4 (23%) | 0 | 0 |
| Improved clinical outcomes | 2 (9%) | 9 (39%) | 12 (52%) | 0 | 0 |
| Helped patient be healthier | 4 (23%) | 15 (65%) | 4 (23%) | 0 | 0 |
| Helped patient be better informed | 11 (49%) | 8 (35%) | 4 (23%) | 0 | 0 |
| Enhanced quality of the visit | 10 (44%) | 9 (39%) | 4 (23%) | 0 | 0 |

The narrative comments that were provided by physicians following the encounters are given in **Table 6** grouped according to major themes. Physicians noted benefits in enhancing patient motivation, providing opportunity to discuss important health issues, improving care efficiency, and offering a professional and personalized approach to discussing fertility and preconception issues. In particular, physicians indicated that the Frame assessment provided the opportunity to candidly discuss sensitive lifestyle topics (weight and obesity, substance use) relevant to reproductive health and helped to prioritize addressing long-standing concerns about irregular cycles that had previously gone unaddressed.

**Table 6.** Physician Narrative Responses Following Frame Discussion with Patients, Organized by Theme.

|  |
| --- |
| **THEME: Patient Motivation or Opportunity** |
| Patient was motivated to discuss preconception and family planning. |
| This patient came to the appointment more motivated than in previous visits, and specifically interested in learning more about WHY she has experienced irregularities in her periods/dysfunctional uterine bleeding. We were finally able to focus our attention on this problem and do a more thorough work-up. Also, she was interested in her family history (which includes sickle cell disease) and understood the importance of preconception visits for testing when she is ready to have a family. She also seems to understand and be more motivated to address these topics. |
| **THEME: Providing an Opening or Opportunity** |
| It was very helpful to go through the patient's complicated psych regimen to help her understand how critical it would be to involve her psychiatrist with changing medications before she becomes pregnant in the future. We were also able to identify some red flags with the patient's menstrual cycle so that she could be aware that coming off birth control in advance of desiring pregnancy and tracking her cycle to ensure ovulation and assess for other diagnoses could be a helpful step. |
| This patient, by participating in Frame, was more ready/willing to discuss her weight (being obese) and to begin to address lifestyle factors for reducing her weight to a healthier value to improve her fertility and future pregnancy outcomes. Prior to this visit she had been reluctant/opposed to such discussions. |
| The Frame evaluation was extremely helpful in evaluating this patient and facilitated a thorough preconception evaluation including ordering lab work and an endometrial biopsy for a problem identified. We were also able to identify some fertility risk factors and provide education on fertility expectations and advice for trying to conceive, as well as lifestyle modifications that will be important to optimize fertility. The patient was extremely appreciative of the thorough care she received. |
| The information provided by Frame helped me to understand my patient's family planning goals so that we were able to have a discussion about PCOS lifestyle modifications to improve fertility, obtained fasting insulin and discussed possibly starting metformin to restore ovulatory function, and we discussed her prior OCP and unwanted side effects so we were able to find an alternative for now while she is not actively trying to conceive, as this could help us treat her iron deficiency (also important prior to pregnancy) and prevent endometrial cancer. |
| **THEME: Clinical Efficiency** |
| It didn't slow the encounter at all, actually helped me understand high priority items and their family planning goals, helped take off a bit of me. |
| I felt like it made me more efficient as it made my conversation more ....efficient, impactful, efficacious |
| This visit was a great example of how Frame can help us to incorporate comprehensive preconception care for eligible women into an annual physical visit. Fertility is something that is not always touched on in an annual physical, but that may be the opportunity for preconception care with many patients. In this patient's case, she had vitamin D deficiency which is now being corrected to optimize fertility, and we were able to discuss optimal timing on how to try to conceive and identify the fertile window, as well as additional labs we could get as part of preconception care – all without adding too much time to her annual visit. |
| **THEME: Personalization** |
| This may seem like a small thing but it is huge that even the assessment "looks pretty and nice" when these patients aren't used to this type of attention to detail and personalization, considering their income and what they can afford in the world of healthcare. Frame is providing care for these Emory patients in a unique and special way, specifically considering that many of them are on Medicaid and typically will just get a packet of papers handed to them on government services. What they are used to is very different from the personalized care that Frame is offering. |

To assess needed inputs for implementation of Frame in primary health care, the clinical coordinators had several observations. They noted that when performing outreach to invite patient participation, many eligible patients reported that they were not planning a pregnancy and, thus, were not interested. When the team further explained that the study was looking at the underlying health of patients of reproductive age, regardless of current or future pregnancy intentions, some who were initially disinterested expressed interest. A summary of lessons learned, based on our study team’s discussion of narrative comments from the clinical coordinator team, is found in **Table 7**.

**Table 7.** Lessons Learned in Implementing Frame Assessment and Discussion in Primary Care.

| **Framing the Preconception Health Risk Assessment and Counseling to Patients** |
| --- |
| Framing the program around reproductive health promotion, rather than pregnancy or fertility planning, is important in helping patients view preconception health risk assessment and counseling as of relevance to them. |
| **Communication with Patients to Arrange Pre-Encounter Completion of Risk Assessment and Coaching** |
| A direct line of communication to the coordinator team, via Google voice and patient portal messaging, is important for timely, direct communication to allow for optimal scheduling of the components of the intervention. |
| An automated way for notifying the coordinator team of patient-initiated changes in scheduled appointments (such as through the electronic medical record system) can contribute to optimal ordering of the components of the intervention. |
| **Prompting Health Care Provider to Access and Review the Preconception Health Risk Assessment** |
| An automated way for notifying the health care provider of the availability of the preconception health risk assessment summary, and in supporting the health care provider to review the summary via a link within the electronic medical record, can contribute to clinical efficiency. |

## Discussion

This feasibility assessment sought to evaluate the integration of preconception health risk assessment and coaching into women’s primary health care encounters at a family medicine clinic. We found that both participating patients, who were individuals assigned female at birth between 21-40 years of age who had a scheduled annual wellness and/or follow-up appointment for a chronic health condition, and their primary health care physicians viewed the process of patients’ completing an online reproductive health risk assessment and virtual coaching session in advance of a primary care encounter in a positive manner. From a patient perspective, the process enhanced confidence in discussing and understanding fertility risks with their health care provider and helped them gain new health information, establish referral to health resources, and motivated health behaviors and actions. Notably, more than half of participating patients reported making lifestyle improvements, nearly a third reported adjusting their birth control and/or prescription medications, and a quarter reported making changes in their planned timing of childbearing. From a physician perspective, the process was viewed as rendering visits more efficient and helping patients be more engaged, better informed and healthier. In particular, physicians noted the process to improve patient motivation and provide opportunity for discussing sensitive lifestyle topics during the primary care encounter.

Additional important findings include that participant sociodemographic and clinical characteristics associated with completion of the virtual coaching and physician discussion of the coaching during the encounter. For completion of the virtual coaching, patient race and ethnicity (non-Hispanic Black patients having lower completion), insurance type (those with government insurance having lower completion), and parity (those with prior births having lower completion) were significantly or nearly significantly associated. Both insurance type and parity were also significantly associated with physician completion of discussion of the assessment, but participant race and ethnicity was not. The reduction in non-Hispanic Black patient completion of the virtual coaching after completion of the digital assessment implies that getting input from Black women into the assessment questions, the assessment and coaching process, and the scheduling of coaching may yield information that would make the Frame process more culturally-congruent or trusted. It is well-known that the cultural tailoring of electronic approaches for the delivery of health interventions is essential for uptake.^17^ That nulliparous patients were more likely to complete the coaching may suggest that they are more concerned about their reproductive health and/or fertility compared to parous patients. That patients covered by government health insurance were less likely to complete the coaching session and have physician discussion at a primary care encounter may suggest that these patients, who generally of lower income with fewer internet, computer, phone, and transportation resources relative to privately-insured patients encountered more barriers. Future studies with low-income primary care populations might involve allowing patients to complete the assessment and schedule the virtual coaching session on-site using a kiosk or tablet rather than in the home setting prior to the scheduled appointment.

The findings of this study contribute to the growing body of literature supporting the feasibility and importance of integrating preconception health promotion into primary care practice. Qualitative studies of female and male primary care patients in Georgia and New York support that both are receptive to the inclusion of reproductive health screening assessments and counseling in primary care practice^18,19^ and that knowledge of preconception health risks improves with brief counseling in the primary care setting.^20^ A cohort study of 300 reproductive aged women in a Canadian primary care practice that implemented a preconception intervention using a three-part model (pre-encounter risk assessment via a tablet, discussion of results with primary care provider, followed by receipt of handouts with risk assessment results and key messages) was effective in identifying preconception health risks, with primary care providers and patients reporting health and practice benefits.^21^ We could not identify studies that have investigated preconception health promotion among gender diverse patients, indicating that studies that include these individuals are needed to inform best approaches.

The study is not without limitations. The study was a small feasibility study and, as such, was of relatively small sample size and did not include longer-term and time-intensive data collection and a comparison group (of similar patients not receiving the intervention) to assess patient behavior and/or health changes over time following the intervention. While the patient sample was diverse with respect to age, race-ethnicity, education and insurance status, it was limited to those assigned female at birth with specific appointment types (wellness exam or follow-up for a chronic health condition).

Our findings support the feasibility of integrating preconception health risk assessment and counseling into primary health care, with important lessons learned. From a market feasibility perspective, our coordinator team’s experience in recruitment and outreach demonstrated that the framing of preconception health risk assessment is important for promoting patient interest. It has been long-recognized that the social marketing of preconception health risk assessment and counseling is challenging given the diverse audience (encompassing all individuals of reproductive age and capability) and a diverse set of health conditions and behaviors.^22^ A particularly challenging group for reaching with preconception health messaging are those who are not currently or in the near future planning to become pregnant.^23^ From a technical feasibility perspective, our experience supports the efficiency in having patients complete the online risk assessment and virtual coaching session but notes that technical tools (such as electronic prompts and embedded links) help the coordination of patient scheduling and the delivery of completed reproductive health risk assessments to the health care provider at the time of the primary care encounter. Given “attrition” from those eligible to those interested among this sample of female primary care patients, it is clear that more feasibility and implementation research is needed to both target the reproductive risk assessment to patients who might not see themselves as being in need of preconception or reproductive health care as well as to understand barriers patients may face in completing online digital assessments. More research is also needed to optimize the workflow for implementing the Frame program into primary care visits, especially for patients with limited access to electronic resources.

## Conclusion

### The problem

Despite recommendations, there is scarce health services research to guide the implementation of preconception health promotion in primary care settings. This feasibility study sought to evaluate the integration of an online preconception health risk assessment and coaching into women’s primary health care at a family medicine clinic.

### Key findings

Female patients and their physicians were highly satisfied with the process of patients’ completing digital risk assessment and counseling prior to the encounter and discussing the assessment during a primary health care encounter. Both patients and physicians viewed the Frame assessment and discussion as being important for improving and/or motivating changes in the patient’s health or health care. Several physicians noted that the process improved patient motivation around lifestyle, improved efficiencies in the clinic, and provided an opening or opportunity for working on lifestyle and reproductive health considerations. To improve uptake, there is room for improving messaging around reproductive health promotion, especially for patients who are not planning to become pregnant in the near future.

### Implications for research and practice

Primary care practices should seek to engage their patients in preconception digital risk assessment and virtual counseling as a means of providing preconception health promotion as recommended by CDC and ACOG. Future research should involve a larger and more diverse sample of primary care patients (to include male and gender diverse individuals) and follow-up over a longer timeframe with more rigorous assessment of longitudinal changes in health behaviors and/or reproductive health outcomes.

## Data Availability

All data produced in the present study are available upon reasonable request to the authors.

